# Circulating tumour cell and cell-free DNA kinetics during radiotherapy in patients with intact head and neck squamous cell carcinoma

**DOI:** 10.1101/2020.10.13.20211516

**Authors:** Sweet Ping Ng, Carolyn S Hall, Salyna Meas, Vanessa N Sarli, Houda Bahig, Carlos E Cardenas, Baher Elgohari, JiHong Wang, Jason M Johnson, Amy C Moreno, Heath Skinner, Adam S Garden, Lumine Na, Ying Yuan, Diana Urbauer, Jack Phan, G Brandon Gunn, Steven J Frank, Shalin Shah, David I Rosenthal, William H Morrison, Michael P MacManus, Clifton D Fuller, Anthony Lucci

## Abstract

Head and neck squamous cell carcinoma (HNSCC) treatment response relies heavily on macroscopic clinical findings. Blood monitoring of circulating markers during treatment may improve earlier detection of responders versus non-responders during radiotherapy. In this study, patients with intact tumour of HNSCC were enrolled in the prospective PREDICT-HN study. Pre-, after first treatment, weekly, and post-treatment blood samples were collected. CTC was enumerated using the CellSearch system. cfDNA was quantified from cfNA isolated at pre-, mid- and post-treatment timepoints. Blood samples were collected from 45 patients. Of the 339 samples analysed for CTC, 31% had detectable CTCs. Nine patients had detectable CTCs (1-3/7.5ml blood) in pre-treatment samples. After 1 fraction, 16 patients had CTCs detected, with 12 who had no pre-treatment CTC. Sixteen (36%) patients had detectable CTC in final week of treatment. There was no correlation between cancer stage, nodal status and tumour burden with CTC. cfDNA levels increased during treatment, with its highest level in the final week and lowest at post-treatment. Our results showed in HNSCC that CTCs can be detected during radiotherapy, suggesting mobilization into circulation during treatment, with as-yet-unknown viability. cfDNA kinetics during treatment correlated with CTC release, and may indicate apoptotic change.

**Simple Summary:** Head and neck squamous cell carcinoma (HNSCC) treatment response relies heavily on macroscopic clinical findings. Blood monitoring of circulating markers such as circulating tumour cell (CTC) and cell-free DNA (cfDNA) during treatment may improve earlier detection of responders versus non-responders during definitive radiotherapy. Although the detection of CTCs and cfDNA in patients with HNSCC has been described, there is minimal data on the kinetics of CTC counts and cfDNA levels during radiotherapy in patients with HNSCC. Here, our study prospectively describes the changes in CTC and cfDNA enumeration during radiotherapy in patients with HNSCC. Our results showed, for the first time to our knowledge, in HNSCC, that CTCs can be detected during radiotherapy, suggesting mobilization into peripheral circulation during treatment, with as-yet-unknown viability. cfDNA kinetics during treatment correlated with CTC release, may indicate apoptotic change during radiotherapy. Combined cfDNA-CTC as an early marker of treatment response should be investigated further.

## 1. Introduction

The annual incidence of head and neck squamous cell carcinoma (HNSCC) in the United States is approximately 50,000 cases, with annual mortality estimated at 11,400 [1]. Radiation therapy is one of the main curative-intent treatment modalities in patients with HNSCC. The current treatment pathway for those who are suitable for definitive radiotherapy treatment is an approximate seven-week course of five daily treatments per week. During radiotherapy, treatment response relies heavily on macroscopic clinical findings and without any overt symptoms suggestive of progressive disease, there are currently no imaging or blood tests that are performed during radiotherapy to stratify responders versus non-responders. Blood monitoring of circulating markers such as circulating tumour cell (CTC) and cell-free DNA (cfDNA) during treatment may improve earlier detection of responders versus non-responders during definitive radiotherapy.

CTCs, first described in 1869, are epithelial cells originating from the tumour, that can be detected within the systemic circulation [2]. These cells are rare and are thought to have evolved from the original malignant tumour, gaining genetic mutations that allow them to enter the systemic circulation, evade and survive the host’s immune system, and exit the circulation and establish a new distant site of disease (seed and soil theory) [3]. Studies have shown the potential use of CTCs as a prognostic and predictive marker in breast [4, 5], prostate [6], gastric [7], colorectal [8], lung [9, 10] and head and neck cancers [9]. cfDNA, on the other hand, are genetic materials found within the systemic circulation that were shed by dying or apoptosed cells [11]. Some of these cfDNA may have genetic characteristics of the malignancy and therefore are classified as circulating tumour DNA [12]. There is early evidence that targeted gene panels may be useful in certain group of patients with HNSCC [13]. As HNSCC is a heterogenous disease, it is a challenge to design a sequence gene panel as a ‘one size fits all’ for all patients with HNSCC with acceptable sensitivity and specificity for detection of minimal residual disease. Therefore, combining cfDNA with other blood markers such as CTC may provide further information and/or improve detection of minimal residual disease in patients with HNSCC.

Although the literature has described the detection of CTCs and cfDNA in patients with HNSCC, there is minimal data on the kinetics or trajectory of CTC counts and cfDNA levels during radiotherapy in patients with HNSCC. Here, our study prospectively describes the changes in CTC and cfDNA enumeration during definitive radiotherapy in patients with HNSCC (PREDICT-HN study) [14].

## 2. Results

This section may be divided by subheadings. It should provide a concise and precise description of the experimental results, their interpretation as well as the experimental conclusions that can be drawn.

### 2.1. Patient characteristics

Overall, 45 patients had completed serial blood collection for analysis. Forty (89%) were males, and the median age of the cohort was 58 years (range: 41 – 81 years). All patients were of good performance status (ECOG 0 – 1). Thirty (67%) patients received photon therapy delivered using volumetric modulated arc therapy (VMAT), and 15 had intensity modulated proton therapy (IMPT). The majority (36 patients, 80%) had concurrent chemotherapy. Table 1 summarizes the cohort’s patient, disease and treatment characteristics.

**Table 1.**
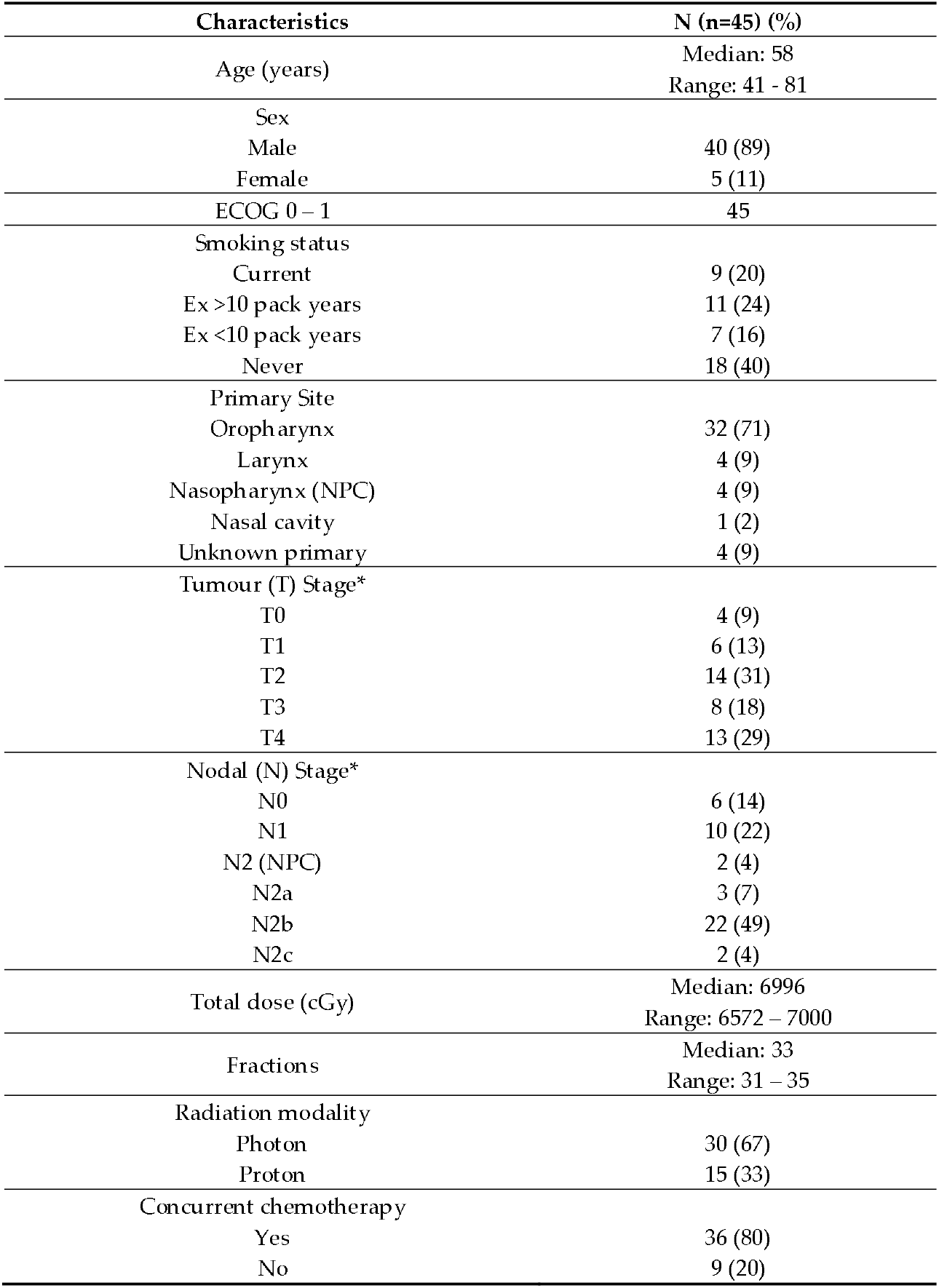
Patients’, tumour and treatment characteristics.

| Characteristics | N (n=45) (%) |
| --- | --- |
| Age (years) | Median: 58<br>Range: 41 - 81 |
| Sex |  |
| Male | 40 (89) |
| Female | 5 (11) |
| ECOG 0 – 1 | 45 |
| Smoking status |  |
| Current | 9 (20) |
| Ex >10 pack years | 11 (24) |
| Ex <10 pack years | 7 (16) |
| Never | 18 (40) |
| Primary Site |  |
| Oropharynx | 32 (71) |
| Larynx | 4 (9) |
| Nasopharynx (NPC) | 4 (9) |
| Nasal cavity | 1 (2) |
| Unknown primary | 4 (9) |
| Tumour (T) Stage* |  |
| T0 | 4 (9) |
| T1 | 6 (13) |
| T2 | 14 (31) |
| T3 | 8 (18) |
| T4 | 13 (29) |
| Nodal (N) Stage* |  |
| N0 | 6 (14) |
| N1 | 10 (22) |
| N2 (NPC) | 2 (4) |
| N2a | 3 (7) |
| N2b | 22 (49) |
| N2c | 2 (4) |
| Total dose (cGy) | Median: 6996<br>Range: 6572 – 7000 |
| Fractions | Median: 33<br>Range: 31 – 35 |
| Radiation modality |  |
| Photon | 30 (67) |
| Proton | 15 (33) |
| Concurrent chemotherapy |  |
| Yes | 36 (80) |
| No | 9 (20) |

### 2.2. Circulating tumour cells (CTC) kinetics

Blood samples were collected at 9 serial timepoints from 45 patients, providing a total of 405 samples. Of those, 22 samples (5.4%) were not analyzed for CTC due to hemolysis and/or unavailability of consumables. Therefore, 383 samples were analyzed. Of the 383 samples, 119 (31%) were positive for CTC.

Nine patients had detectable CTCs (1 – 3 CTCs per 7.5ml blood) in pre-treatment samples. After 1 fraction of radiotherapy, 16 patients had CTCs detected (1- 5 CTCs per 7.5ml blood), of which 12 patients had no CTC detected pre-treatment. Figure 1 illustrates the overall kinetics of CTC counts during radiotherapy for this cohort of patients. All patients had detectable CTC at some point during radiotherapy except for 3 patients who had no CTC detected pre, during and post-treatment. Sixteen (36%) patients had detectable CTC in the final week of treatment. Eleven patients had CTC detected 2 to 3 months post-treatment. All 11 patients did not have detectable CTC pre-treatment. Due to the small number of CTCs detected (1 – 5 CTCs), no significant change of quantity of CTCs between timepoints was detected.

**Figure 1.**
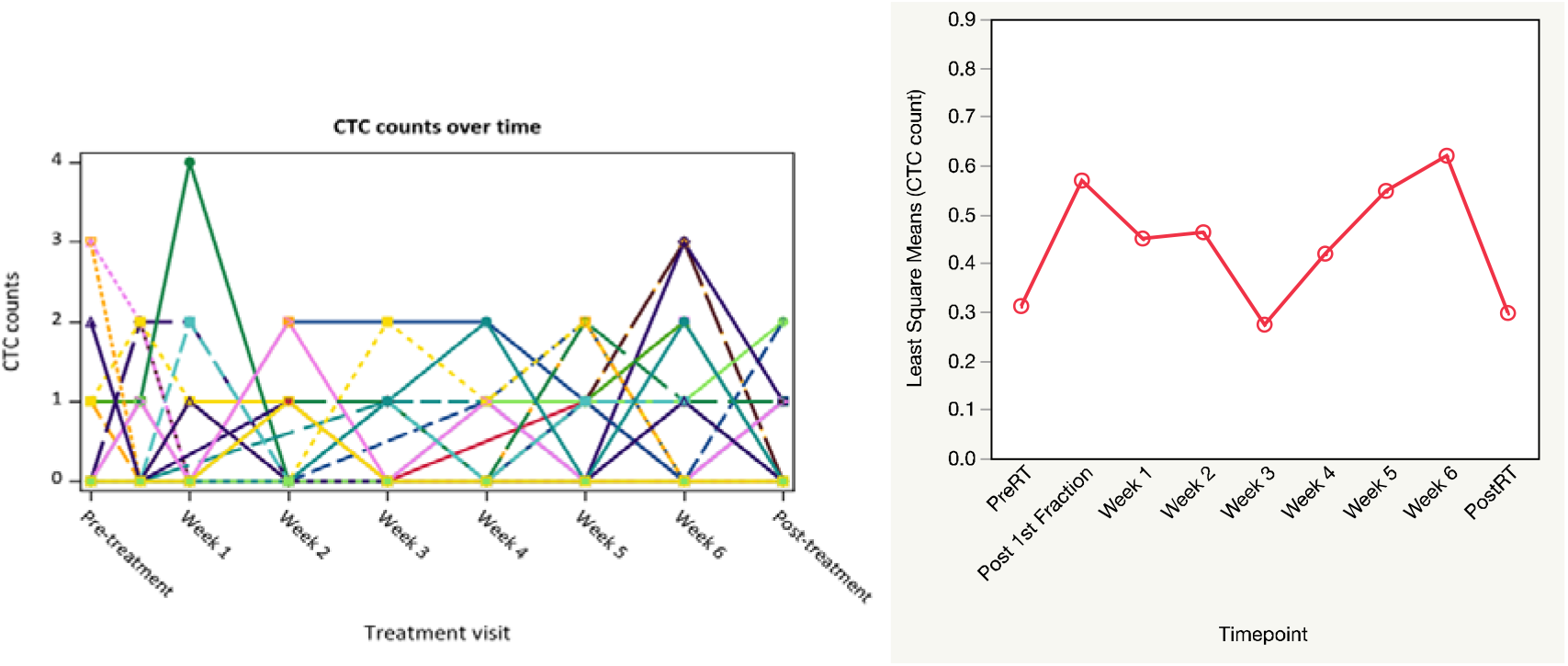
This is a figure, Schemes follow the same formatting. Kinetics of circulating tumour cell (CTC) counts during radiotherapy.

### 2.3. CTC and GTV correlation

Out of 45 patients, four patients did not complete the MRI component of the study. Therefore, 41 patients had serial weekly MR images – 36 had primary tumour evaluable, and 31 had nodal tumour evaluated. No significant correlation was detected between primary GTV size and CTC count for each timepoint (Table 2). CTC count correlated with nodal tumour volume in week 4, but no significant correlation was observed at other timepoints (Table 2).

**Table 2.**
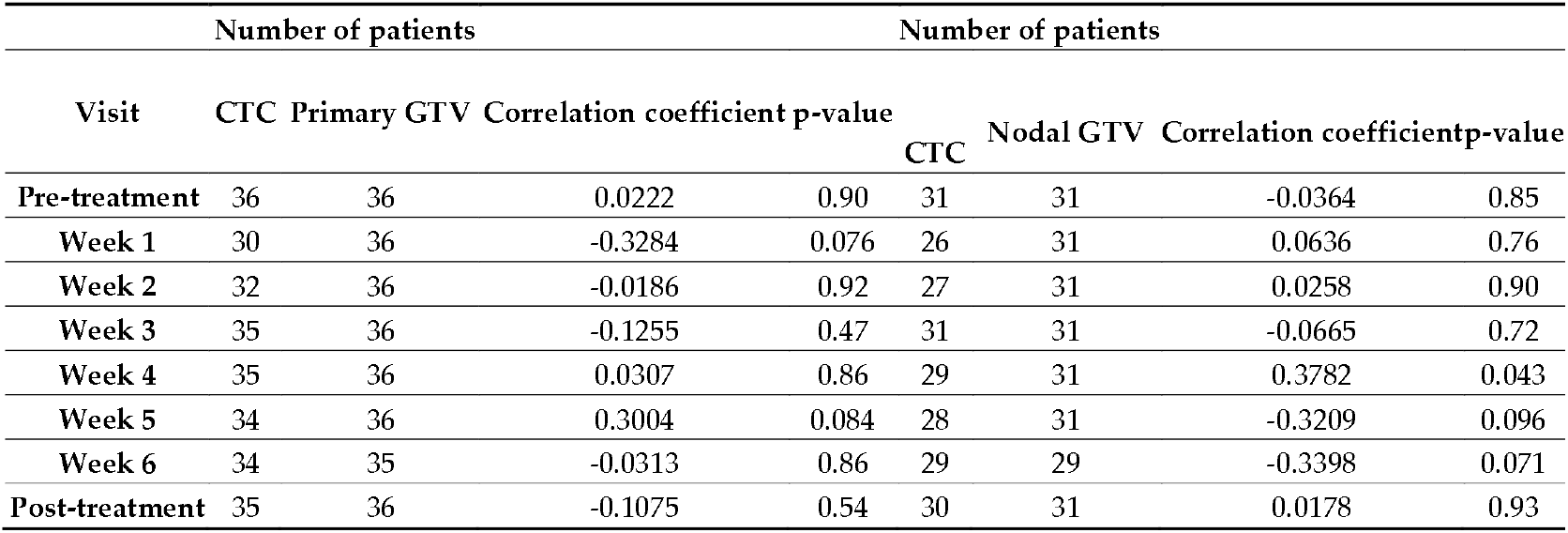
Correlation between CTC counts and primary and nodal gross tumour volume (GTV).

### 2.4. CTC and cfDNA kinetics

Overall the circulating cfDNA levels increased during treatment, with its highest level in the final week of treatment, and lowest at post-treatment timepoints. As depicted in Figure 2, the levels of circulating cfDNA remained relatively stable between pre-treatment and mid-treatment (week 4) timepoints. As the number of CTCs increase in weeks 3 to 6, a rise in cfDNA was detected between mid-treatment and end-treatment (week 6) timepoints.

**Figure 2.**
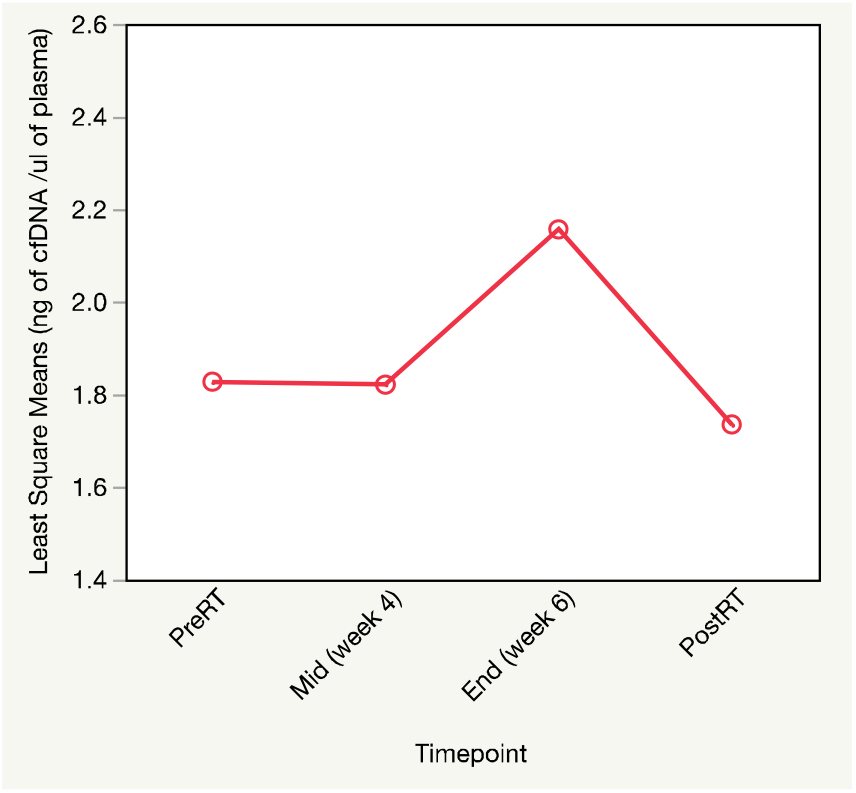
Kinetics of total circulating cell-free DNA (cfDNA) levels during radiotherapy.

### 2.5. CTC and outcomes

The median follow-up was 12 months (range: 3 – 22 months). The 18-month overall survival and progression free survival were 94% and 81%, respectively (Figure 3). Eight patients developed disease recurrences – 2 with local disease, 2 with regional disease, 2 with local and regional disease, and 2 with distant disease. The CTC kinetics for these patients are depicted in Supplemental Figure 1. Two patients (both with local and regional recurrences) died with disease during this period. With limited number of events, further correlative statistical analysis was not performed.

**Figure 3.**
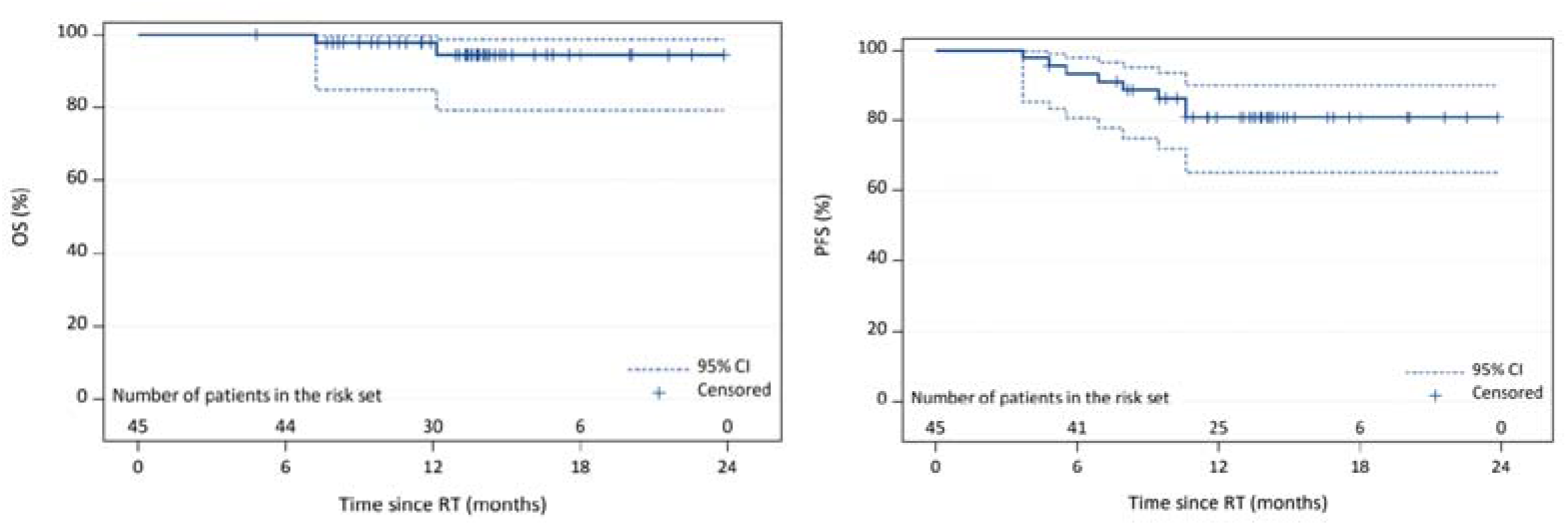
Overall survival and progression free survival for the cohort.

## 3. Discussion

Our study showed that there was an increase in CTC numbers after first fraction of radiotherapy, and from mid-treatment (week 3) to end of radiotherapy, in patients undergoing definitive radiotherapy for head and neck squamous cell carcinoma. Furthermore, we observed that total cell-free DNA also increased from mid-treatment to end of radiotherapy.

Our study demonstrated that the trajectory of CTC counts was not linear, and did not correspond with tumour shrinkage during radiotherapy in head and neck cancer. This highlights the importance of an initial systematic assessment of the kinetics of CTC during treatment, rather than pre-selecting timepoints for convenience assuming a linear response or reduction in number of CTC to treatment, in future studies for any other tumour sites. In fact, we showed that the number of CTCs rise after a single fraction of radiotherapy, indicating mobilization of CTC into the systemic circulation by radiation. Our findings are consistent with a study by Martin et al [17] in patients with lung cancer, where they detected an increase in the number of CTCs in patients who received radiotherapy for non-small cell lung cancer. Their group performed immunostaining of detected CTCs with γ-H2AX antibodies and the CTCs demonstrated high γ-H2AX signal indicating that these cells were from the irradiated tumour site. The notion that local cancer therapy, such as surgery and radiotherapy, can potentially mobilize tumour cells into the systemic circulation and potentially influence the risk of development of distant metastatic disease, is not novel. Several surgical studies have reported an increase in CTC counts during and/or after oncological surgery in patients [18-23]. With regards to radiotherapy, although not demonstrated in human studies until more recently, Kaplan and Murphy [24], in 1949, showed that lung metastases were more common in tumour-bearing mice that received radiotherapy than those were not irradiated. However, the significance and impact of CTC release into the systemic circulation by oncological treatments on patients’ outcomes remains unknown.

Although we observed a drop in CTC counts from 1^st^ fraction of radiotherapy to week 3, we noted a rise in the CTC counts in the latter half of the radiotherapy course. The viability and metastatic potential of these CTCs is unknown. However, Martin et al [17] previously demonstrated viability of isolated CTCs after radiotherapy from patients with lung cancer, suggesting that CTCs mobilized by radiotherapy have the capacity to ‘seed and soil’ and establish at a distant site. Furthermore, we also observed a rise in total cfDNA from mid-treatment to end of treatment. As cfDNA are DNA fragments released into the circulation as a result of cell death or apoptosis, the rise in cfDNA levels coinciding with the increase in CTC numbers towards the end of treatment may suggest that the CTC detected were dead or dying cells. Pleasingly, we observed that the levels of CTCs and cfDNA dropped from final week of treatment to post-treatment timepoint, potentially indicating successful eradication of disease in some patients, as the detection of either CTC or cfDNA could potentially indicate residual disease within the patient [25]. Longer follow up is required in our cohort to determine the predictive value of detected residual CTC and cfDNA at the post-treatment timepoint. Given the heterogenous group of patients and low level of cfDNA, we did not proceed to quantify the circulating tumour cfDNA which would confirm if the circulating DNA originated from the tumour. Previous studies have indicated difficulty in validating results of tumour cfDNA in patients with head and neck cancer due to the small number of patients, heterogeneity of tumour sites and stages, and lack of agreement of molecular targets to quantify [25, 26]. Regardless, this could potentially be investigated in our cohort in the future, once there is further validation and standardization of molecular targets.

This study comes with its limitations. Firstly, although prospectively evaluated, we had a small cohort of patients. This study was designed to assess feasibility of collecting serial paired MR imaging and blood from patients undergoing radiotherapy for head and neck cancer. At the time of study design, there was limited evidence on the detectability and enumeration of CTC during radiotherapy in patients with localized head and neck cancer. With a small cohort of patients, statistical analysis was limited. Secondly, our study included a heterogeneous group of patients with HNSCC of different mucosal primaries and of different tumour (T) and nodal (N) stages. However, we demonstrated that CTC can be detected during radiotherapy regardless of primary site of disease, and disease stage. The lack of correlation between CTC and GTV may indicate that these two parameters provide different independent information of the disease biology. Longer follow up is required to demonstrate if either is predictive of subsequent outcomes. Thirdly, we must acknowledge the limitations of using CellSearch method to enumerate CTCs in our study. Being an EpCAM-based method of detecting CTC, this method detects malignant cells of epithelial origin such as squamous cell, but cancer cells that have undergone epithelial-to-mesenchymal transition (EMT) may not be detected. During EMT, to optimize metastatic potential, malignant cells lose their epithelial markers and gain mesenchymal qualities such as vimentin instead [27, 28]. Furthermore, the viability of the CTC released into the circulation, particularly after first fraction of radiotherapy, cannot be determined as cells captured by CellSearch are permeabilized and cannot be isolated and cultured [16]. Another challenge in using CellSearch for CTC quantification is that the number of CTCs detected is reportedly lower than other methods of CTC isolation and quantification [17, 29, 30]. Nevertheless, now that we have preliminary data that CTCs are released into the circulation during radiotherapy, other methods to capture CTCs for culture to determine viability and metastatic potential can be investigated.

## 4. Materials and Methods

### 4.1. Patient selection

All eligible patients seen at The University of Texas MD Anderson Cancer Center from May 2017 through August 2018 were offered participation in this prospective observational PREDICT-HN study approved by the institutional review board (ClinicalTrials.gov Identifier: NCT03491176). The full study protocol has been previously published [14]. Eligibility criteria include pathologic diagnosis of squamous cell carcinoma, intact primary and/or nodal tumour visible on imaging, and good performance status (ECOG 0 – 1). Patients with evidence of distant metastatic disease, had contraindications to MR imaging, or had previous head and neck irradiation less than 5 years ago were excluded. Informed consent was obtained from the participant prior to any study-related imaging or blood collection.

### 4.2. Radiotherapy treatment

Patients had definitive radiotherapy as per institutional standard of care protocol, after pathological diagnosis and complete staging work up. All cases were discussed at the institutional head and neck tumour board prior to treatment. Patients were treated with curative-intent radiotherapy with/ without concurrent chemotherapy. All patients were reviewed, discussed and their radiation therapy contours reviewed at the departmental head and neck quality assurance clinic by head and neck radiation oncologists.

### 4.3. Blood collection and circulating tumour cell (CTC) enumeration

Whole blood was collected aseptically by venipuncture or from a venous port into a 10ml CellSave preservation tube and two 10ml EDTA tubes at the specified serial timepoints (Figure 1). The presence and quantification of CTCs were performed using blood collected in the CellSave tube using the standardized, semiautomatic FDA-approved Cellsearch system. The CellSearch method of CTC enumeration was previously described in detail [4, 15, 16]. In brief, the blood-filled CellSave tube was centrifuged to separate plasma from solid blood components. Following immunomagnetic capture and enrichment of cells expressing epithelial cell adhesion molecule (EpCAM)-with antibody-coated nanoparticles from 7.5ml of blood, these cells were stained with fluorescent nucleic acid dye 4,2-diaminodino-2-phenylindole hydrochlroride (DAPI), and antibodies specific for epithelial cells (cytokeratin 8,18, 19 – phycoerythrin) and leukocytes (CD45-allophycocyanin). A semiautomated fluorescence-based microscope system was used to identify CTCs. CTCs were defined as cytokeratin positive and CD-45 negative nucleated cells [4]. A qualified laboratory technician reviewed and confirmed all results.

All blood collected in CellSave tubes were analysed for CTCs within 72 hours of collection. To avoid intervention-related CTC release into circulation, the pre-radiotherapy blood was collected at least a week after any invasive procedure, such as a biopsy.

### 4.4. cfDNA enumeration

Blood collected in the EDTA tubes were centrifuged and the plasma was filtered and collected prior to storage at -80C. For cfDNA enumeration, 4ml of frozen plasma from pre-, mid (week 4), end (week 6) and post-radiotherapy were used. cfNA was isolated using the commercially available MagMAX Nucleic Acid Isolation Kit (Thermo Fisher Scientific). cfDNA was quantified using Qubit high sensitivity dsDNA assay (Invitrogen).

### 4.5. Magnetic resonance imaging (MRI)

Patients had pre-treatment, weekly during treatment, and post-treatment MRI scans. Pre-treatment MRI was obtained within 2 weeks before commencement of radiotherapy. Patients were imaged in radiotherapy treatment position, and immobilized with a thermoplastic mask using the MAGNETOM Aera 1.5T MR scanner (Siemens Healthcare, Germany) with two 4-channel large flex phased-array coils, and 32-channel phased-array spine coil. Specific MR sequences and parameters were detailed in the published protocol [14].

### 4.6. Statistical analysis

Missing values were not imputed: where there were missing data for an outcome variable, those records were removed from any formal statistical analysis of that outcome variable (complete case analysis), unless otherwise specified. Graphs and descriptive statistics were used to understand characteristics of GTV and CTC, and relationship between GTV and CTC. The correlation between CTC count and the gross tumour volume (GTV) was calculated using Spearman’s rank correlation coefficient. The linear mixed model (LMM) was used to evaluate change in GTV, CTCs and cfDNA over time. Factors associated with post-treatment outcomes (GTV and CTC) change from pre-treatment were examined using linear regression adjusted for pre-treatment value, unless otherwise specified.

The time to events (overall survival and progression free survival) were described using Kaplan-Meier methods. The time to events were estimated using Kaplan-Meier method and time to events outcome estimate at months 6, 12, and 18 were provided with 95% confidence intervals. All analyses and data manipulations were carried out using SAS 9.4 and JMP 14.0 (SAS Institute Inc.).

## 5. Conclusions

In conclusion, our results showed, for the first time to our knowledge, in head and neck squamous cell carcinoma, that CTCs can be detected during radiotherapy, suggesting mobilization into peripheral circulation during treatment, with as-yet-unknown viability. cfDNA kinetics during treatment correlated with CTC release, may (and hopefully) indicate apoptotic change during radiotherapy. Combined cfDNA-CTC as an early marker of treatment response should be prospectively and systematically investigated.

## Data Availability

Data are embargoed pending peer-review and data descriptor publication; in the interim, data are available upon request at the discretion of PI(s) via contact email. A meta-data link for public data deposition is included for post-embargo data deposition.

https://figshare.com/articles/dataset/_/13082309

## Supplementary Materials

The following are available online at www.mdpi.com/xxx/s1, Figure S1: Circulating tumour cell (CTC) counts for patients who developed disease recurrence

## Author Contributions

Conceptualization, S.P.N., C.S.H., M.P.M., C.D.F., and A.L.; methodology, S.P.N., C.S.H., M.P.M., C.D.F., and A.L.; software, C.E.C., and C.S.H; validation, S.M., J.M.J, V.N.S, and H.B.; formal analysis, S.P.N., L.N., Y.Y., and D.U..; investigation, S.P.N.; resources, S.P.N., J.W., J.M.J., A.M., H.S., A.S.G., G.B.G., J.P., S.J.F., D.I.R., W.H.M., and S.S.; data curation, S.P.N., S.M, V.N.S., H.B., C.E.C., and B.E.; writing—original draft preparation, S.P.N.; writing—review and editing, S.P.N., J.W., J.M.J., A.M., H.S., A.S.G., G.B.G., J.P., S.J.F., D.I.R., W.H.M., C.S.H., M.P.M., C.D.F., A.L., L.N., Y.Y., D.U., S.M, V.N.S., H.B., C.E.C., and B.E; visualization, S.P.N.; supervision, C.D.F., C.S.H., M.P.M., and A.L.; project administration, S.P.N, C.S.H., C.D.F., M.P.M., and A.L.; funding acquisition, S.P.N, C.D.F, and A.L. All authors have read and agreed to the published version of the manuscript.

